# Beyond Hormones: The Efficacy of Time-Restricted Eating as an Alternative to Traditional Pharmacological Therapy for Polycystic Ovary Syndrome

**DOI:** 10.1101/2025.05.21.25327203

**Authors:** Isabela von Damm, Marcela Hernández-Ortega, Cynthia Dickter Sarfati, Federico W von Son

## Abstract

**Background/Objective:** Polycystic ovary syndrome (PCOS) is one of the most common endocrinopathies during reproductive age, affecting up to 25% of women worldwide. The aim of this study was to evaluate the efficacy of Time Restricted Eating (TRE) as an alternative to traditional hormonal therapy in patients with PCOS.

**Methods:** A prospective, longitudinal, comparative clinical study was conducted. A total of 16 patients, diagnosed with PCOS, were divided into three groups to assess combined and isolated treatment options. Two groups underwent a 90-day Time Restricted Eating protocol with measurements at day 0 and 90 for ovary follicle count, luteinizing hormone, follicle stimulating Hormone, estradiol, progesterone, total testosterone, free testosterone, fasting insulin and glucose. Results were compared to a control group undergoing traditional hormonal treatment.

**Results:** Ultrasonographic assessment of left and right ovary, showed a significant reduction in FC for the TRE group (36.67 ± 12.81 % and 41.79 ± 3.76, respectively). Metabolic and hormonal markers also showed an important reduction: HOMA-IR (11.7 ± 8.47 %), GLU (5.06 ± 0.91 %), INS (11.87 ± 6.93 %), FT (54.92 ± 10.01%), TT (3.96 ± 1.48%) LH (22.67 ± 3.24%) and FSH (23.53 ± 6.84 %).

**Conclusions:** A 90-day TRE intervention is a safe and reliable treatment option when compared to standard Oral Contraceptive (OC) therapy, especially for the improvement of hyperandrogenism, metabolic and hormonal parameters in patients with PCOS.

## INTRODUCTION

Polycystic ovary syndrome (PCOS) is one of the most common endocrinopathies during reproductive age, affecting up to 26% of women worldwide ^[1,2]^. The most common signs and symptoms include, anovulatory and/or absent menstrual cycles, hyperandrogenism and its concomitant signs, among others. Additionally, one of the main concerns, due to the menstrual irregularities, is that fertility can be compromised during the course of the disease. In a recent systematic review, Yuqi Liu et al demonstrated that improving insulin sensitivity increase the chances of pregnancy for patients with PCOS ^[3]^.

Even though PCOS’ pathophysiology has been described as multifactorial, its association with insulin resistance and its effect on hyperandrogenism has been widely studied ^[4].^ This condition is present in 23-35%^[5–7]^ of PCOS cases, where insulin sensitivity reduces approximately 35-40% ^[4,7]^, leading to a compensatory overproduction of insulin (INS).

Given that insulin acts directly on the ovaries increasing the frequency of luteinizing hormone (LH) pulses, hyperinsulinemia overstimulates theca cells, producing androgens in a constant and higher rate, which can intervene with the hypothalamic-pituitary-ovary axis (HPOA), contributing to anovulation and the formation of cysts^[8]^. On the other hand, these high levels of insulin act on the liver inhibiting sex hormone binding protein (SHBP) synthesis ^[4,9,10]^. This decrease in SHBP leads to higher concentrations of free testosterone (FT), causing acne, hirsutism, hypertrichosis, virilization and alopecia, as described in figure 1.

**Figure 1.**
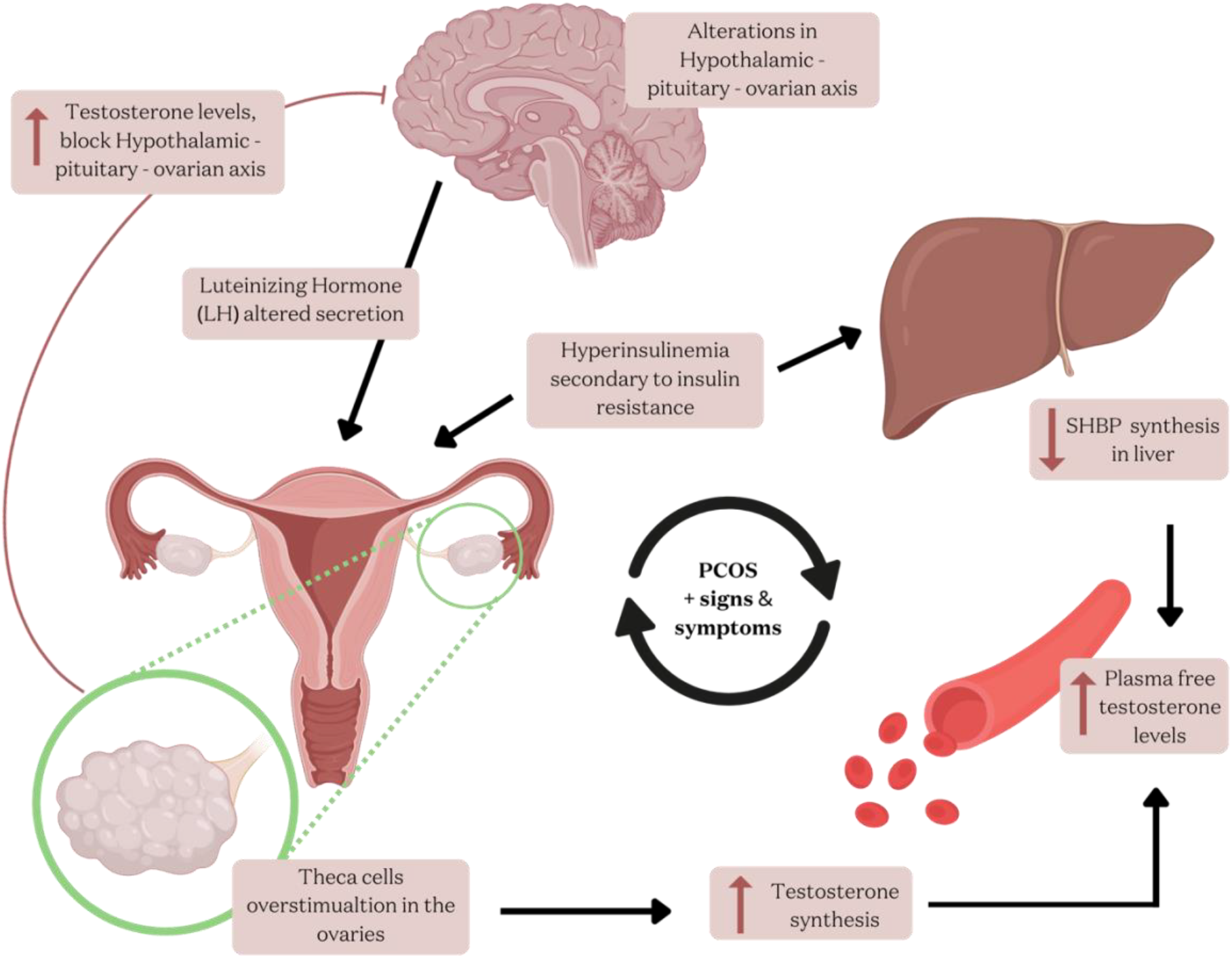
Altered glucose and insulin metabolism induce hyperinsulinemia, which consequently acts in the ovarian theca cells causing an overproduction of androgens. High levels of plasmatic free testosterone add up to the neuro endocrine axis imbalance. (modified from Subbulaxmi Trikudanathan, MD, MRCP, MMSc Polycystic Ovarian Syndrome. Med Clin N Am 99. 2015; pp. 221–235.)

According to international guidelines, the standard treatment for insulin resistance-related PCOS combines oral contraceptive (OC) therapy^[11]^, with insulin sensitizers, such as metformin ^[12,13]^ The use of OC inhibits the natural secretion of LH, follicle stimulating hormone (FSH), estrogen and progesterone, which can have long term negative side effects^[14,15]^. Due to the estrogenic component of OC, patients have a two-fold increase risk of venous thromboembolism, and risk of developing breast cancer increases up to 30% ^[14,16,17]^. These severe side effects, oblige clinicians and researchers to study novel, safer and sustainable therapeutic approaches for a diagnosis that threatens women’s life quality.

Intermittent fasting (IF) and time restricted eating (TRE) have gained relevant popularity in the last decades. The term IF encompasses different strategies that vary on time of exposure to calories and circadian rhythmicity. This generally produces metabolic improvements in humans, impacting in the reduction of cardiovascular risk, insulin, cholesterol, triglycerides and low density lipoprotein (LDL) levels, ^[18–20]^. First described by Panda et al in mice ^[21]^ and afterwards tested in humans ^[22]^, 16 hours of fasting followed by 8 hours of ad-libitum calorie intake (16:8) on non-consecutive days, is one of the most commonly practiced and adaptable ways of TRE. Other strategies use 18 or 20 hours of fasted periods with 6 and 4 hour eating windows (18:6, 20:4), respectively.

Multiple studies have demonstrated that IF corrects glucose homeostasis by improving insulin signaling ^[23,24].^ A physiological positive loop could be found between TRE interventions and PCOS hormonal pathways, through the increase in insulin sensitivity, which in turn could result in the downregulation of androgen overproduction ^[25]^.

This study aims to evaluate the effect of an 18:6 fasting intervention to reduce the hyperandrogenic signs and symptoms and potentially reverse the diagnosis of PCOS.

## MATERIALS AND METHODS

### Study design

Approval from the research and ethics committee at ABC Medical Centre, Mexico City, was obtained on 04/19/2021, (registration number: ABC-21-14), This research respected the guidelines of the Declaration of Helsinki and complied with the guidelines established in NOM-012-SSA3-2012 from México. The study was also registered under ClinicalTrials.gov (NCT06031753).

### Participants

Patient enrollment was conducted through social media, using mainly Instagram stories, Instagram posts and live sessions to explain the impact of the protocol. Patients preregistered through an online google-forms questionnaire and if they met the initial criteria, applicants were then contacted by the research team for final interview and assessment. If eligible, patients received orientation and explanation of the scope of the clinical study and signed a consent form to join the protocol.

After diagnosis of PCOS, according to Rotterdam criteria^[26]^ the following parameters were considered to determine eligibility: (1) age (20-35), moderate physical activity, body mass index (BMI) 20-24.9 kg/m^2^, (2) active treatment with OC for group CP and MIX, nulliparity, “A, B or C” PCOS phenotypes ^[27–29].^ Exclusion criteria were: (1) post or perimenopause, “D” PCOS phenotype, (2) night shifts, (3) comorbidities, (4) additional medical treatment and (5) having conducted any form of IF previously. Additionally patients were eliminated if they reported frequent or excessive consumption of alcohol or any drugs.

Subject to eligibility criteria, a total of 30 participants were recruited. Twenty pre-screened patients (undergoing active OC treatment) were randomized for groups CP and MIX, the 10 remaining participants who were not taking OC, were automatically assigned to group IF. Dropouts modified the final cohort to 16, due to the following reasons: pregnancy during the intervention (n = 1), not returning for last control (n = 8), unwilling to adhere to TRE intervention (n = 4), diagnosis of autoimmune disease (n = 1).

Group CP, considered as the control group, included participants with previous diagnosis of PCOS undergoing standard treatment with combined oral contraceptives (ethinyl estradiol + progestin), without additional changes in nutritional, lifestyle and/or exercise variables. Group MIX were participants with previous diagnosis of PCOS undergoing standard treatment with oral contraceptives, which started a TRE protocol with an eating window of 6 hours and a fasting window of 18 hours (18:6). Group IF included participants with previous diagnosis of PCOS without additional medical treatment, who started an 18:6 fasting regime. It is very important to outline that group IF, had never used any previous medication, such as oral contraceptives and/or any others, for the treatment or management of PCOS. The research group performing ultrasonographic evaluation and sample collection was blinded and patients were randomly assigned to each member.

### Clinical determination of PCOS and follow up

A transvaginal transductor was used to perform ultrasound (equipment: GE logiq 5 expert), considering measurements of ovarian volume, as well as volume and number of follicles. Hormonal levels of estrogen, progesterone, LH, FSH, total testosterone (TT) and free testosterone were measured. Fasted blood samples were collected and clinical assessment of hyperandrogenism was evaluated through anamnesis. Rotterdam criteria were used to determine PCOS phenotypes (A,B,C,D), through the aforementioned markers.

### Metabolic and hormonal assessment

A baseline control using venous blood samples, was taken at day 0 with a follow up testing at day 90, which included: LH, FSH, estradiol, progesterone, TT, FT, fasting INS and glucose (GLU), with a minimum of 8 hour fasting. Fasting INS was measured using an Architect C16200 equipment, through chemiluminescence technique. Fasting glucose was determined through automated spectrophotometry with an Architect C16200 equipment. For the remaining hormones (LH, FSH, prolactin, estradiol, progesterone and total testosterone) a chemiluminescence technique was implemented, using an Architect i4000 equipment. Finally free testosterone was measured through ELISA essay using STATFAX 4700.

### Determination of insulin sensitivity

To determine the insulin sensitivity status of the participants, the HOMA – IR equation was applied, as determined by Matthews et al ^[30]^. This value represents a clinical indicator calculated through fasting insulin and glucose levels, which uses the following formula:

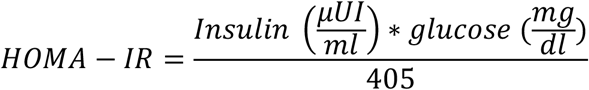

HOMA-IR was calculated after obtaining values of each sample collection, at baseline and day 90.

### Statistical analysis

All data were analyzed with GraphPad Prism v.10.1.1. ^[31].^ The percentage of change (reduction or increase) of each of the variables obtained from each of the patients with respect to the initial values was calculated, using the following formula:

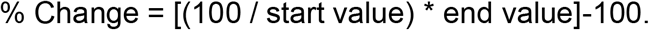

The individual values obtained were grouped by treatment, participants with the highest and lowest value in each variable were not considered for the analysis to avoid outlier bias, and the mean and standard error were obtained. These averages between the three treatment groups were analyzed using different tests based on the nature of the data.

## Results

Final patient distribution for data analysis considered 6-5-5 for groups CP, MIX and IF, respectively. As expected, metabolic improvement was noted in the intervention groups, with relevant modifications in glucose homeostasis and insulin sensitivity. Control group showed an important increase in insulin levels, with a very low change in glucose and HOMA-IR values.

Interestingly, the use of oral contraceptives (group CP) showed an increase in insulin levels and HOMA-IR with respect to initial values, by 27.36 ± 11.0% and 10.93 ± 9.12%, respectively. The combined intervention (MIX) showed a reduction of 20.28 ± 2.84% for insulin and 24.05 ± 4.36% for HOMA-IR. Finally, patients subject to exclusive Intermittent Fasting (IF) treatment reflected a reduction of 11.87 ± 6.93% in insulin levels and 11.7 ± 8.47% for HOMA-IR (Mean ± SE). Highly significant differences were found between the CP and MIX groups for insulin (P=0.014) and HOMA-IR (P=0.024) and significant differences in CP vs IF (P=0.034) for insulin, please refer to figure 2.

**Figure 2.**
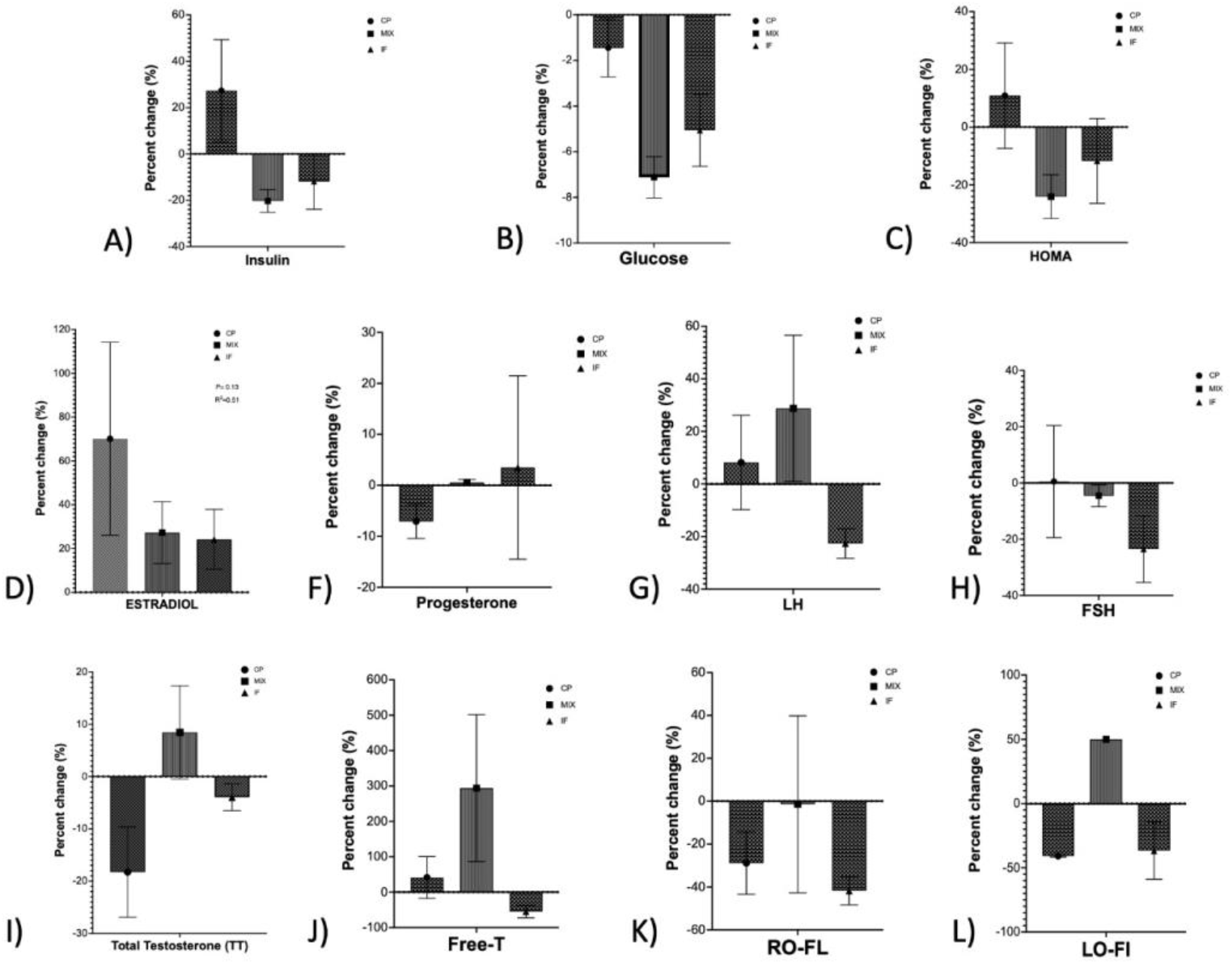
changes in metabolic, hormonal markers and follicle count in right (RO-FC) and left (LO-FC) ovaries. CP: control group with oral contraceptive treatment, without TRE intervention. MIX: combined group with oral contraceptive treatment and TRE intervention. IF: Time restricted eating intervention group without hormonal treatment. RO-FC: right ovary follicle count. LO-FC: left ovary follicle count.

For glucose parameters, all groups showed a reduction when compared to baseline, with percentage changes of 1.45 ± 0.63%, 7.12 ± 0.52% and 5.06 ± 0.91% (Mean ± SE) for groups CP, MIX and IF, correspondingly. Highly significant differences were found between the CP and MIX groups (P=0.001) and significant differences in CP vs IF groups (P=0.033).

Surprisingly, within the hormonal panels, FT and TT revealed an important increase in the MIX group (294.00 ± 119.9 % and 8.45 ± 5.16 %). On the other hand, in the group treated only with TRE (group IF) both testosterones showed an important decrease of 3.96 ± 1.48% for TT, and 54.92 ± 10.01% (Mean ± SE) for FT. However, in the CP group, FT and TT exhibited opposite behavior; TT decreased by 18.26 ± 4.31%, while FT increased by 41.45 ± 29.5 %. Statistically significant differences were only evident when comparing FT levels in CP vs IF (P=0.041) and MIX vs IF (P=0.025).

No statistically significant differences were found between any of the treatments (p=0.13) when assessing estradiol levels. With an average increase of 70.12 ± 22.06%, 27.28 ± 8.17% and 24.19 ± 7.91% for CP, MIX and IF, respectively (Mean ± SE). Additionally, LH values increased in the CP group by 8.18% ± 8.99%. The MIX group showed an increase in LH and progesterone, with a decrease in FSH levels (28.76 ± 16.06 %, 0.57 ± 0.33 % and 4.55 ± 2.23 %, respectively). Lastly, patients in the IF group had an increase in progesterone by 3.5 ± 10.39%, although a decrease in LH (22.67 ± 3.24%) and FSH by 23.53 ± 6.84 % (Mean ± SE) was observed. Only statistically significant differences between FSH percent change were found when comparing MIX and IF (0=0.048). Luteinizing hormone showed statistical significance only for PC vs IF (P=0.035) and progesterone for PC vs MIX (P=0.049).

All treatments showed a reduction in the number of follicles found in the right ovary (RO-FC), with reductions of 28.89 ± 7.28%, 1.43 ± 29.19%, 41.79 ± 3.76, for the CP, MIX and IF treatments, respectively (Mean ± SE). No statistically significant differences were found between groups (P=0.49). Only the CP and IF treatments showed a reduction in the quantity of follicles in the left ovary (LO-FC) of 40.83 ± 0.57 % and 36.67 ± 12.81 %, respectively (Mean ± SE); the mixed treatment group reported an increase of 50%. Highly significant differences were found between CP vs MIX (P=0.004) and statistically significant differences between IF and MIX groups (P=0.021).

## Discussion

The efficacy of OC in treating PCOS is arguable, since pharmacological treatment’s side effects seem to outweigh its benefits when compared to placebo ^[32]^ and it has been described that women can even develop post-pill syndrome ^[33].^ Unfortunately, the use of OC, is a palliative approach that focuses in signs and symptoms temporarily, compromising physiological ovary function and provoking anovulation and fertility disorders ^[34,35]^, in addition to the popular and known side effects which can impair women’s health^[35]^. This being said, it should become imperative to explore and include lifestyle interventions when treating this diagnosis. Intermittent fasting has shown to be an effective strategy for metabolic control and hormone regulation, nevertheless, research performed in women with PCOS has its limitations and we ought to be critical with its scope and benefits ^[36]^.

Some studies evaluating the effects of fasting during Ramadan ^[37]^, have shown some slight, positive outcomes on metabolic parameters and ovary function. Nevertheless, caution should be taken when interpreting these results, since it is well understood that TRE and circadian rhythmicity are highly correlated and could alter hormonal status and, consequently, the final results ^[21,22]^. Furthermore, this study is a retrospective analysis, which didn’t include a control group, nor an oral contraceptive comparison cohort.

In a study conducted by Rothschild et al ^[38]^ a decrease in 70% of HOMA-IR was observed after an 18 week TRE intervention. In the present research, intermittent fasting reduced INS levels in both intervention groups by up to 24.05 ± 4.36 %. On the other hand, our control group had an increment in fasted insulin levels, which guides our conclusions towards thinking that conventional treatment can deteriorate metabolic status, as shown in previous studies^[39]^, potentially disguising an ovarian dysfunction. The notable decrease in INS in both groups undergoing TRE, as a consequence ameliorated HOMA-IR, which shows an improved insulin signaling and sensitization. Once INS dysfunction resolves, hence, glucose homeostasis is achieved (as shown in the drop of glucose levels in the intervention groups), various positive, quantifiable responses are present in PCOS patients.

C. Li et al. conducted a TRE trial in 15 patients with PCOS, but exhibited multiple limitations ^[40]^. First of all, the duration of the study was only 5 weeks and they performed a 1 week weight stabilization phase, which could have produced additional bias in some parameters. Compared to our study, C. Li et al, analyzed data of overweight and obese women; we, on the other hand, excluded these groups to avoid confounding variables. However, we observed similar results in decreases of hormonal markers, such as LH, TT, FSH and INS.

The hepatic response to the reduced insulin levels is to increase the SHBP production and secretion, which in turn, induces the reverse transport of a larger amount of FT for its degradation in the liver, decreasing its plasma levels, thus, the hyperandrogenic signs and symptoms. We also hypothesize that the decrease in FT in our intervention group correlates with the increase in SHBP observed in the C. Li trial.

In the ovaries, diminution of LH regulates the theca cell overstimulation, provoking as a result a decrease in androgen production. In patients with active PCOS, increased LH levels cause anovulation and follicle retention (retained follicles cause the pathognomonic ultrasonographic image, also described as polycystic ovary). In our IF group, both ovaries showed notable decrease in follicle number with reductions of 41.79 ± 3.76 % in the right ovary and 36.67 ± 12.81 % for the left. Hong et al demonstrated that follicle count (FC) in patients with PCOS is highly correlated with metabolic dysfunction and IR (^[41]^). This data supports our findings and explains the imminent connection between IR and PCOS. Our outcome is a novel finding that demonstrates the impact of regulating INS and IR through TRE to improve PCOS ovary’s phenotype.

Additionally, the important drop of FSH observed in the IF group, could be linked to a reduction in the overstimulation provoked by high levels of FT in the hypothalamus. Downregulation of FT levels creates a positive loop that could require less FSH (as observed in our results) to carry out ovary function through LH. These findings suggest that patients can regulate their neuro-endocrine axis activity with TRE interventions, thus resulting in positive changes in the ovary function & fertility.

Surprisingly, in the MIX group, we observed unexpected outcomes. Both testosterone values increased when compared to IF and OC and hormones from the ovarian axis showed contrary behaviors, LH increased by 28.76 ± 16.06 % and FSH decreased by 4.55 ± 2.23 %. Additionally both ovaries showed an increase in follicle count, suggesting that combining fasting strategies with oral contraceptive treatment in PCOS doesn’t seem to exhibit a positive response from a gynecological perspective. Nonetheless, metabolic improvement was achieved, with proper response to INS, GLU and HOMA-IR.

Finally, we observed that the OC group, as expected, revealed a positive change in FC and TT, in concordance with international guidelines. The positive change in the formation of cyst in the ovaries is a result of the arrest in the natural hormonal cycle elicited by the administration of synthetic hormones. Meaning, that OC therapy does not promote physiological hormonal fluctuations, essential for fertility, ovulation and PCOS resolution.

We also demonstrated that patients undergoing conventional treatment worsened their metabolic profiles, which according to our study aggravates the dysfunctional axis between INS and the ovary, failing to resolve the underlying causes of insulin resistant PCOS.

## Conclusion

To our knowledge, this is the first study that demonstrates an improvement in PCOS phenotypes, in both metabolic and hormonal health, using a novel and safe lifestyle intervention as an eighteen hour TRE protocol during 90 days. As shown in our results, fasting has demonstrated to be a promising substitute for PCOS standard hormonal treatment, avoiding the downsides and negative side effects of oral contraceptives. However, we suggest that further research is warranted in the field of endocrine-gynecology and fasting interventions for different phenotypes and pathologies.

## Data Availability

All data produced in the present study are available upon reasonable request to the authors

## Acknowledgements

We want to thank all the participants who committed and completed our study to contribute in this groundbreaking research that can potentially improve the life of many women. The authors acknowledge Dr. Elke von Son for assistance provided on the statistical analysis. We also gratefully acknowledge the intervention provided by Dr. José Eduardo Serratos and all the staff and lab members.

